# Effects of Integrated Traditional Chinese and Western Medicine on Knee Joint Functional Stability Reconstruction Following Anterior Cruciate Ligament Reconstruction : protocol for a assessor-blinded, randomized controlled trial

**DOI:** 10.1101/2025.03.27.25324809

**Authors:** Xinglai Zhang, Chen Chen, Guo Lu, Hang Gao, Jiayi Ren, Yuanjia Gu, Shaohua Chen, Jiming Tao

## Abstract

**Introduction:** Anterior Cruciate Ligament Reconstruction (ACLR) imposes substantial socioeconomic and healthcare burdens worldwide. In China, Traditional Chinese Medicine (TCM) has emerged as a popular complementary and alternative medicine strategy to relive pain and functional stability. However, the current evidence is insufficient to support the efficacy of TCM in addressing knee pain and improving physical function. This trial aims to assess the clinical effectiveness of TCM.

**Methods and analysis:** A total of 132 patients, aged between 18 and 50 years, will be recruited from Shuguang hospital. Eligible participants will be randomly allocated to either the experimental group receiving integrated TCM-Western rehabilitation intervention five sessions or the control group receiving conventional rehabilitation intervention per week. Both groups will undergo a 4-week intervention phase, followed by a 4-week follow-up period. The primary outcome measure is the change from baseline in Lysholm Score, Knee Muscle Strength, the Knee Injury and Osteoarthritis Outcome Score (KOOS), effusion, Muscle Thickness, The modified Star Excursion Balance Test (SEBT), Knee Range of Motion (ROM), Knee Proprioception. All adverse events occurring during the trial will be promptly documented.

**Ethics and dissemination:** This study protocol has been approved by the Ethics Committee of Shuguang Hospital Affiliated to Shanghai University of Traditional Chinese Medicine (Approval Number: 2024-1522-105-01). Prior to enrollment, written informed consent will be obtained from all eligible participants in accordance with ethical guidelines. Upon study completion, findings will be disseminated through peer-reviewed publications in academic journals and presented at relevant scientific conferences to ensure broad knowledge dissemination and contribute to evidence-based clinical practice.

**Trial registration:** The trial was registered on the Chinese Clinical Trial Platform (Registration No. ChiCTR2400085239)

**Funding:** Key Supporting Discipline Construction Projects in Shanghai’s Health System (2023ZDFC0301)

## Introduction

The rapid socioeconomic development in China has been accompanied by increased participation in outdoor activities, rising incidence of traffic accidents, population aging, and lifestyle transitions—all contributing to a significant upsurge in knee injuries. Among these, anterior cruciate ligament (ACL) ruptures and meniscal tears rank as the most prevalent traumatic knee injuries, with international studies reporting an ACL injury rate of 1 in every 3,500 individuals and approximately 154 meniscal injuries per 100,000 people annually [1-4]. Clinically, autologous tendon graft reconstruction for ACL tears and surgical repair/partial resection for meniscal damage are standard interventions to restore joint biomechanical stability [5]. However, emerging evidence highlights substantial limitations: up to two-thirds of patients fail to regain pre-injury athletic performance within one-year post-ACLR, while nearly a quarter experience secondary ACL injury [6-7]. These outcomes underscore a critical disparity—surgical procedures effectively repair structural damage but cannot restore neuromotor control stability, leaving unresolved rehabilitation challenges such as persistent pain and swelling [9], quadriceps weakness [10], restricted range of motion [11], proprioceptive deficits [12], postural instability [13], gait abnormalities [14], central nervous system plasticity alterations [15], and diminished quality of life [16].

The 2022 Evidence-Based Clinical Guidelines for ACL Injury Management advocate for early standardized rehabilitation emphasizing immediate post-ACLR knee mobilization and closed-chain exercises [17]. Yet, global consensus on optimal rehabilitation protocols remains elusive [18]. Conventional programs, structured around tissue healing timelines, prioritize pain management, passive knee extension, quadriceps strengthening, early mobilization, partial weight-bearing (under complication-free gait conditions), and functional training [19]. Disappointingly, these approaches yield re-injury rates as high as 30% post-return-to-sport [20], underscoring the urgent need for innovative, evidence-based rehabilitation strategies that bridge the neuromotor recovery gap.

Emerging evidence from prior research suggests the therapeutic potential of Traditional Chinese Medicine (TCM) in augmenting ACLR recovery. Specifically, early postoperative Tuina massage along the Spleen, Stomach, and Gallbladder meridians, combined with acupressure at key acupoints including Yanglingquan (GB34), Fenglong (ST40), Xuanzhong (GB39), Zusanli (ST36), and Chongyang (ST42), has demonstrated improvements in knee range of motion (ROM), swelling reduction, and functional scores [21]. Adjunctive acupuncture at Zusanli (ST36), Yanglingquan (GB34), and Yinlingquan (SP9) further alleviates post-arthroscopic pain, edema, and functional impairments [22]. Notably, ultra-early integration of TCM therapies with rehabilitation training significantly enhances pain management, swelling control, ROM restoration, and functional outcomes at the 3-month milestone [23]. Additionally, early TCM intervention improves proprioceptive recovery and functional restoration [24]. While existing literature highlights TCM’s early-phase benefits in ACLR recovery, no studies have systematically evaluated its long-term impacts on neuromuscular stability—a critical determinant of re-injury risk and sustained functional recovery[25]. Given the absence of standardized TCM protocols for ACLR rehabilitation, this study proposes to develop and validate an evidence-based, integrated TCM-Western rehabilitation framework. Through a randomized controlled trial (RCT), we will compare the efficacy of this integrated protocol against conventional care across multiple domains: knee functional scores, pain/swelling indices, muscle strength/morphometry, balance/proprioception, ROM, limb symmetry, gait biomechanics, and central nervous system adaptations. Comprehensive assessments at short-term and mid-term follow-ups will evaluate TCM’s modulatory effects on knee joint functional stability.

## Patients and Methods

### Study design and settings

This investigation was conducted as a prospective, parallel group randomized controlled trial (RCT) with repeated measures design. A total of 132 postoperative anterior cruciate ligament reconstruction (ACLR) patients were enrolled and allocated in a 1:1 ratio to either the experimental group or the control group using stratified randomization. Participants in the experimental cohort received an integrated TCM-Western rehabilitation intervention, while those in the control cohort followed conventional rehabilitation protocols. The study duration spanned 8 weeks, consisting of a 4-week active treatment phase immediately followed by a 4-week follow-up observation period. This structured design allowed for longitudinal assessment of intervention effects across both short-term recovery and early-phase functional stabilization.

### Participants and Recruitment

The diagnosis of anterior cruciate ligament (ACL) injury was based on current medical history, physical examination, and MRI-related criteria outlined in Practical Orthopedics [26].This study recruits participants who have undergone anterior cruciate ligament reconstruction (ACLR) from both outpatient clinics and inpatient wards of the Rehabilitation Department at Shuguang Hospital. All participants must meet the following criteria:

### Inclusion criteria

1. Confirmed diagnosis of anterior cruciate ligament (ACL) injury and isolated arthroscopic ACL reconstruction (ACLR) within the preceding month. Preoperative full functional capacity of the affected knee, with postoperative subjective reports of instability.
2. Adults aged 18–50 years (inclusive), regardless of gender.
3. No history of trauma, surgery, or clinical instability in the non-operated knee.
4. Minimum baseline Tegner Activity Scale score of 5
5. No documented history of hypersensitivity to herbal medications or related supplements.
6. Full understanding of the rehabilitation protocol and provision of written informed consent prior to enrollment.

### Exclusion criteria

1. Presence of multiple ligament ruptures or trauma affecting joints other than the reconstructed knee
2. New fractures, nonunion, or delayed union within the past 3 months. Unrepaired meniscal tears of Grade II or higher (based on MRI or arthroscopic evaluation) or chondral lesions identified during preoperative assessment.
3. History of trauma or surgery involving the lumbar spine, hip, knee, or ankle. Active rheumatoid arthritis, severe osteoporosis, or lower extremity deep vein thrombosis.
4. Documented cognitive impairment. Uncontrolled psychiatric conditions or inability to comprehend/comply with study protocols due to emotional instability or communication barriers.
5. Active, severe primary diseases of the cardiovascular, hepatic, renal, or hematological systems.

### Sample size calculation

Based on prior research and the sample size estimation formula for comparing two means of continuous data, we calculated the required sample size using the Lysholm Knee Score as the primary outcome measure. According to published literature [27], we assumed a mean score (μ_2_) of 44.8 in the experimental group and 41.7 (μ_1_) in the control group, with a standard deviation (σ) of 5.0 and a minimum clinically important difference (μ_2_ - μ_1_) of 3.1. Setting α (type I error) at 0.05 and β (type II error) at 0.10 (90% power), the initial calculation yielded a sample size of 55 participants per group. To account for a 20% dropout rate, we inflated the sample size to 66 participants per group, resulting in a total recruitment target of 132 subjects.

### Randomization and blinding

Participants with anterior cruciate ligament reconstruction (ACLR) were recruited from outpatient clinics at the Rehabilitation Medicine Department of Shuguang Hospital, Shanghai University of Traditional Chinese Medicine. Eligible patients meeting inclusion/exclusion criteria were randomized 1:1 to experimental or control groups using SPSS 22.0-generated sequences by an independent statistician. Allocation concealment was ensured via sequentially numbered, opaque envelopes containing group assignments, with patient screening IDs on the exterior. Envelopes were opened post-consent, immediately before intervention assignment, adhering to CONSORT guidelines to minimize selection bias.

Due to intervention nature, therapists and participants could not be blinded. However, outcome assessors, statisticians, data collectors, and administrators remained blinded throughout the study. Blinding was strictly enforced, with codes held by an independent party and broken only post-analysis to ensure unbiased interpretation. This method complies with CONSORT standards for transparent blinding in RCTs.

### Intervention

Prior to trial commencement, all physicians and therapists will undergo standardized operating procedure training to ensure full comprehension of trial protocols. Participants are discouraged from pursuing additional interventions during the trial period to maintain intervention integrity. While medications may be administered if medically necessary, their type and dosage will be meticulously documented. To facilitate participant compliance and access, all treatment-related costs will be covered by the study, ensuring no financial burden on participants.

### Control group

The conventional rehabilitation program was designed in accordance with the Evidence-Based Clinical Guidelines for Anterior Cruciate Ligament Injury (2022 Edition) [17] and relevant international guidelines for postoperative ACL management [25]. It encompassed knee range-of-motion exercises, knee and hip strength training, balance training, and functional drills. All rehabilitation sessions were conducted by therapists certified by the National Health Commission of the People’s Republic of China with over three years of clinical experience. The intervention was administered once daily, five days per week, for a total duration of four weeks. Detailed treatment parameters are followed. Rehabilitation schedule for weeks 1-2:①Patellar mobilization: 2 sets of 10 repetitions. ②Heel slides with knee flexion (0-90°): 3 sets of 10 repetitions. ③ Isometric quadriceps contractions. ④ Straight leg raises (SLR): 3 sets of 10 repetitions. ⑤Gluteal bridges: 3 sets of 10 repetitions. ⑥Ambulation with normal gait using crutches. ⑦Ankle resistance exercises. ⑧Hip flexion, extension, abduction, and adduction training. Rehabilitation schedule for weeks 3-4: In addition to the above protocol. ①Increase knee flexion angle, achieving full range of motion by 6 weeks postoperatively. ② Gradual reduction of crutch dependency, progressing to walking without orthotics or crutches. ③ Partial squats (knee flexion 0-60°): 3 sets of 10 repetitions. ④Step-up training. ⑤Vastus medialis obliquus strengthening exercises. ⑥Stationary cycling: 15 minutes.

### Experimental group

In addition to conventional rehabilitation, participants received an integrated traditional Chinese medicine (TCM) intervention package comprising acupuncture, Tuina (Chinese therapeutic massage), and herbal cold compresses. ①Tuina: Patients were positioned supine. The affected lower limb was treated with one-finger Zen pushing, finger kneading, and pushing maneuvers along the Spleen Meridian of the Foot (SP), Stomach Meridian of the Foot (ST), and Gallbladder Meridian of the Foot (GB), applied proximally from the ankle to the thigh. Each technique was performed for five cycles with gentle pressure, administered once daily. ② Acupoint Stimulation: Acupoints Yongquan (KI1), Jiexi (ST41), Chongyang (ST42), Sanyinjiao (SP6), Yinlingquan (SP9), Xuanzhong (GB39), Zusanli (ST36), Yanglingquan (GB34), Fenglong (ST40), Xuehai (SP10), Liangqiu (ST34), Futu (ST32), Fengshi (GB31), Chongmen (SP12), Chengshan (BL57), and Weizhong (BL40) were pressed. Each point was stimulated for 10 seconds after achieving de qi (arrival of qi), administered once daily. ③Herbal Cold Compress: A TCM herbal formula was prepared by decocting 1000 mL of water down to 300 mL, then cooled to 0°C–5°C. The cooled solution was applied as a cold compress to the affected knee using thick gauze pads, twice daily (morning and afternoon). The herbal formula comprised Rhei Radix et Rhizoma (Dahuang) 50 g, Coptidis Rhizoma (Huanglian) 50 g, Phellodendri Chinensis Cortex (Huangbai) 50 g, and Scutellariae Radix (Huangqin) 50 g. ④Acupuncture: Acupuncture was performed using a draining (reducing) technique at meridians-based points and local points around the knee. Specific points included Shenshu (BL23), Guanyuan (CV4), Dubi (ST35), Xiyan (EX-LE4), Liangqiu (ST34), Xuehai (SP10), Yanglingquan (GB34), and Xiyangguan (GB33), administered once daily.

TCM therapies were delivered by therapists and acupuncturists certified by the National Health Commission of the People’s Republic of China with over three years of clinical experience. The intervention was administered five days per week for a total duration of four weeks.

### Outcome Measures

#### Primary Outcome Measures

##### Lysholm Score

Primarily utilized for knee ligament injuries, this score is characterized by high accuracy and specificity. It evaluates eight dimensions: limping, locking, pain, stair climbing, support use, instability, swelling, and squatting, with a total score of 100 points. The score incorporates both subjective patient-reported functional assessments and objective clinical signs. Lysholm scores were recorded pre-intervention, at week 4 post-intervention, and at week 8 post-intervention.

#### Secondary Outcome Measures

##### Knee Muscle Strength

Isokinetic knee extension and flexion muscle strength were measured using the HUMAC NORM Isokinetic Dynamometer System (CSMI, USA). Following five maximal-effort warm-up repetitions and a 3-minute rest period, participants performed five maximal-effort knee extensions and flexions at angular velocities of 60°/s and 120°/s, respectively, and ten maximal-effort repetitions at 300°/s, with 20 seconds of rest between each velocity. Measurements were taken for both limbs, and peak torque values for the quadriceps and hamstrings of both lower extremities, as well as the H/Q ratio, were recorded. Isokinetic knee strength measurements were recorded at week 4 and week 8 post-intervention.

##### Pain

Pain was assessed using the Knee Injury and Osteoarthritis Outcome Score (KOOS) pain subscale. This scale ranges from 0 (extreme pain) to 100 (no pain), with higher scores indicating reduced pain. Pain scores were recorded pre-intervention, at week 4 post-intervention, and at week 8 post-intervention.

##### Effusion

Participants lay supine, and knee circumference at the midpoint of the patella was measured using a flexible tape measure to the nearest 0.1 cm to assess effusion.

The average of three repeated measurements was calculated. The intra-class correlation coefficient (ICC), standard error of measurement (SEM), and minimal detectable change (MDC) were 0.97, 0.04 cm, and 0.2 cm, respectively. Knee circumference measurements at the midpoint of the patella were recorded pre-intervention, at week 4 post-intervention, and at week 8 post-intervention.

##### Muscle Thickness

Muscle thickness was measured using the Apogee 1000 color Doppler ultrasound system. Participants lay supine with their hands naturally placed by their sides. The lower limbs were fully positioned on the bed, and participants relaxed for 5 minutes prior to measurement. With the thigh skin fully exposed and lower limb muscles completely relaxed, the ultrasound probe was placed at the midpoint of the line connecting the anterior superior iliac spine and the upper part of the patella, parallel to the line and perpendicular to the surface of the rectus femoris. The probe was coupled with the skin without compressing the soft tissue. The probe angle was adjusted slightly until the ultrasound image was clear, with distinct muscle and fascia layers, facilitating marking and measurement. The ultrasound detection depth was then adjusted to ensure that the rectus femoris, intermediate femoral muscle, and femur were within the ultrasound image range. The specific contour ranges of the rectus femoris and intermediate femoral muscle were identified, marked with points, and connected with vertical measurement lines. Muscle thickness values for the rectus femoris and intermediate femoral muscle were obtained based on the machine’s measurements. Muscle thickness measurements for the rectus femoris and intermediate femoral muscle were recorded pre-intervention, at week 4 post-intervention, and at week 8 post-intervention.

##### Balance Function

The modified Star Excursion Balance Test (SEBT) was used to assess dynamic postural control of the lower limbs in anterior, posteromedial, and posterolateral directions. Participants performed a light jogging warm-up at a self-selected pace for 5 minutes and six familiarization trials in each reach direction prior to testing. Three attempts were made in each direction, and measurements were recorded to the nearest 0.5 cm. All distance scores were normalized to leg length (%LL). SEBT results were recorded pre-intervention, at week 4 post-intervention, and at week 8 post-intervention.

##### Knee Range of Motion (ROM)

Knee ROM (°) was assessed using a goniometer, with participants lying supine and their heels elevated on a foam roller. For knee extension measurements, participants maximally extended their knees, and the difference from 0° extension was defined. For knee flexion measurements, participants bent their knees and slid their heels towards their buttocks as far as possible. Side-to-side difference scores were calculated as follows: Difference (°) = (Uninjured -Injured). Knee ROM measurements were recorded pre-intervention, at week 4 post-intervention, and at week 8 post-intervention.

##### Knee Proprioception

The CSMI Isokinetic Dynamometer System was used to assess passive and active knee joint position sense. Absolute angular error (AAE) was calculated. Proprioception results were recorded at week 4 and week 8 post-intervention. Gait Biomechanics and Surface Electromyography (sEMG): Three-dimensional gait acquisition and analysis were performed using the ONONATE 3D motion capture gait analysis system (Shanghai Maiwo Medical Technology Co., Ltd.). Participants were instructed to walk at a self-selected speed for 20 meters, and knee, hip, and ankle angles, as well as plantar pressure distribution, were recorded. sEMG signals were collected synchronously with the 3D gait analysis system. Eight muscles on both sides, including the rectus femoris, biceps femoris, tibialis anterior, and gastrocnemius, were selected. Raw sEMG signals from the relevant muscles were recorded synchronously during 3D gait analysis and automatically transmitted from the control terminal to the computer, where they were saved in the 3D gait analysis database. sEMG signals were smoothed, rectified, band-pass filtered (10-500 Hz), and time normalized. Data included raw data, root mean square amplitude (RMS), activation period (ACTI), integrated EMG, and median frequency. Gait analysis results were recorded at week 4 and week 8 post-intervention.

## Adverse events

This study implements a structured adverse event (AE) management system, emphasizing ethical compliance and participant safety. AEs, defined as post-treatment medical occurrences (e.g., pain, cardiovascular symptoms), are documented via standardized protocols in Case Report Forms (CRFs), including CTCAE severity grading and causality assessments. A tiered response mechanism guides evaluation: clinicians report suspected AEs, investigators apply protocol-driven workflows, and final adjudication integrates subjective/objective data. Severe AEs (SAEs), such as life-threatening events, require expedited 24-hour ethics committee reporting, with emergency communication for urgent cases. Multidimensional data collection and dynamic risk assessment ensure trial integrity and participant welfare, balancing safety with scientific rigor.

### Statistical analysis

Statistical analyses were performed using IBM SPSS Statistics Version 26.0. Descriptive statistics were presented as percentages for categorical variables and means ± standard deviations (Mean±SD) for continuous variables, with 95% confidence intervals (CIs). Independent-samples t-tests were applied to continuous dependent variables, while Fisher’s exact tests evaluated categorical data (gender, graft type, dominant/affected limb). Normality was verified via Shapiro-Wilks tests (p>0.05), and homogeneity of variances (if applicable) was assessed using Levene’s tests (p>0.05). For repeated-measures ANOVA violating sphericity assumptions (Mauchly’s test), Greenhouse-Geisser corrections were reported. A 2 × 3 (group × time) mixed-model ANOVA examined interaction effects between the within-subjects factor “time” (pre-intervention, week 4, week 8) and between-subjects factor “group” (experimental vs. control). Significant interactions prompted Benjamini-Hochberg-corrected post-hoc paired t-tests. Isokinetic strength across velocities was analyzed using 2 × 2 (group × time) repeated-measures ANOVA. Significance was set at p<0.05. Effect sizes were interpreted using Cohen’s d: weak <0.2, weak-to-moderate 0.2–0.4, moderate 0.4–0.65, moderate-to-strong 0.65–0.7, strong >0.8.

## Data management

Screeners will collect comprehensive baseline demographic and clinical data at the time of patient enrollment. Outcome assessments will be documented in standardized Case Report Forms (CRFs) by trained evaluators. Two independent data administrators, external to the investigative team and blinded to group allocations, will undergo rigorous training in data management protocols prior to engaging in double-data entry using Microsoft Excel. Real-time data will be uploaded to the Chinese Clinical Trial Registration Center’s electronic data capture system, enabling continuous monitoring by the Science and Technology Department at Shuguang Hospital. This system ensures compliance with regulatory standards through automated data validation checks and audit trails, maintaining data integrity throughout the trial lifecycle.

## Quality control

Under the oversight of a dedicated steering committee, rigorous quality control measures will be systematically implemented across all trial phases. Prior to study initiation, investigators will complete mandatory training in standardized trial methodologies and longitudinal monitoring protocols to ensure procedural consistency. Any protocol modifications or corrective actions will be formally documented and submitted to both the steering committee and institutional ethics committee for approval, adhering to established regulatory frameworks. This structured governance model ensures compliance with scientific integrity standards while maintaining operational transparency throughout the study conduct.

## Discussion

Patients who have undergone anterior cruciate ligament reconstruction (ACLR) often experience a variety of symptoms, including pain, swelling, decreased proprioception, and impaired walking function, which significantly hinder their return to family and work. Traditional Chinese therapies such as Tuina, acupuncture, and herbal medicine are deeply trusted and welcomed by patients.

Tuina, through its manual manipulation of specific body surface areas, effectively regulates the physiological and pathological states of the body, promotes local blood circulation, and alleviates muscle tension and pain. For postoperative patients, Tuina can enhance blood circulation in the soft tissues around the knee joint, accelerate the resolution of swelling, and increase joint mobility[28]. Acupoint stimulation, on the other hand, modulates qi and blood flow as well as visceral functions by stimulating specific acupoints. For instance, acupoints such as Zusanli (ST36) and Yanglingquan (GB34) have the effects of invigorating the spleen and replenishing qi, promoting blood circulation, and activating meridians, which are beneficial for alleviating postoperative pain and promoting the recovery of knee joint function[29].

As a core therapy in TCM, acupuncture achieves therapeutic purposes by regulating meridian qi and blood flow and dredging meridians. Studies have shown that acupuncture can improve local qi and blood circulation in the knee joint, reduce inflammatory responses, and promote tissue repair. When combined with rehabilitation training, acupuncture significantly enhances postoperative knee joint function scores and shortens recovery time[30]. Herbal cold compress, made from herbs with heat-clearing, detoxifying, and blood-activating properties, can reduce swelling and pain, promote blood circulation, and alleviate postoperative swelling and pain when applied to the affected area, thereby facilitating the recovery of knee joint function.

Numerous studies have confirmed that the combination of TCM and conventional training can significantly improve knee joint function in postoperative patients[21,22]. Tuina combined with rehabilitation training can enhance knee joint mobility and reduce joint stiffness, while acupuncture combined with rehabilitation training significantly improves functional scores and quality of life. Furthermore, TCM therapies have shown remarkable efficacy in alleviating postoperative pain. Acupoint stimulation and acupuncture exert analgesic effects by regulating the nervous system, while herbal cold compress alleviates pain by reducing local temperature and mitigating inflammatory responses.

TCM therapies can also promote postoperative tissue repair and regeneration. Acupuncture stimulates local blood circulation, increases nutrient supply to tissues, and accelerates the repair process. The blood-activating and stasis-resolving components in herbal cold compress also promote tissue repair. TCM therapies are characterized by high safety, minimal side effects, good patient tolerability, and easy operation, making them suitable for clinical promotion. Additionally, TCM treatment plans can be individualized according to the specific conditions of patients, meeting their diverse needs.

Despite the promising clinical outcomes of TCM combined with conventional training, further optimization of treatment protocols is necessary. In the future, the exploration of optimal combinations of different TCM methods and their best synergy with conventional training may enhance therapeutic efficacy. Currently, the mechanisms of action of TCM combined with conventional training are not fully understood. Further research is needed to investigate these mechanisms at the molecular, cellular, and tissue levels to provide a solid theoretical foundation for clinical applications.

In conclusion, the integration of TCM with conventional training demonstrates remarkable efficacy in the rehabilitation of patients following ACLR, offering multiple advantages such as improved knee joint function, pain alleviation, and tissue repair promotion. Its safety and feasibility make it highly valuable for clinical promotion. Future research should focus on optimizing treatment protocols, elucidating the mechanisms of action, and conducting large-scale, multicenter studies to further validate its clinical efficacy and safety.

## Data Availability

no data

## Trial status

At the time of submission, recruitment for the trial has been started. The first participant was included on 5 August 2025. The study is scheduled to end in December 2027.

## Acknowledgements

The authors would like to thank all the investigators, study staff, outcome assessors, participants and their families for their support in this trial.

## Patient and public involvement

Patients and/or the public were involved in the design, or conduct, or reporting, or dissemination plans of this research. Refer to the Methods section for further details.

## Patient consent

for publication Consent obtained directly from patient(s).

## Provenance and peer review

Not commissioned; externally peer reviewed.

---

**Table 1.**
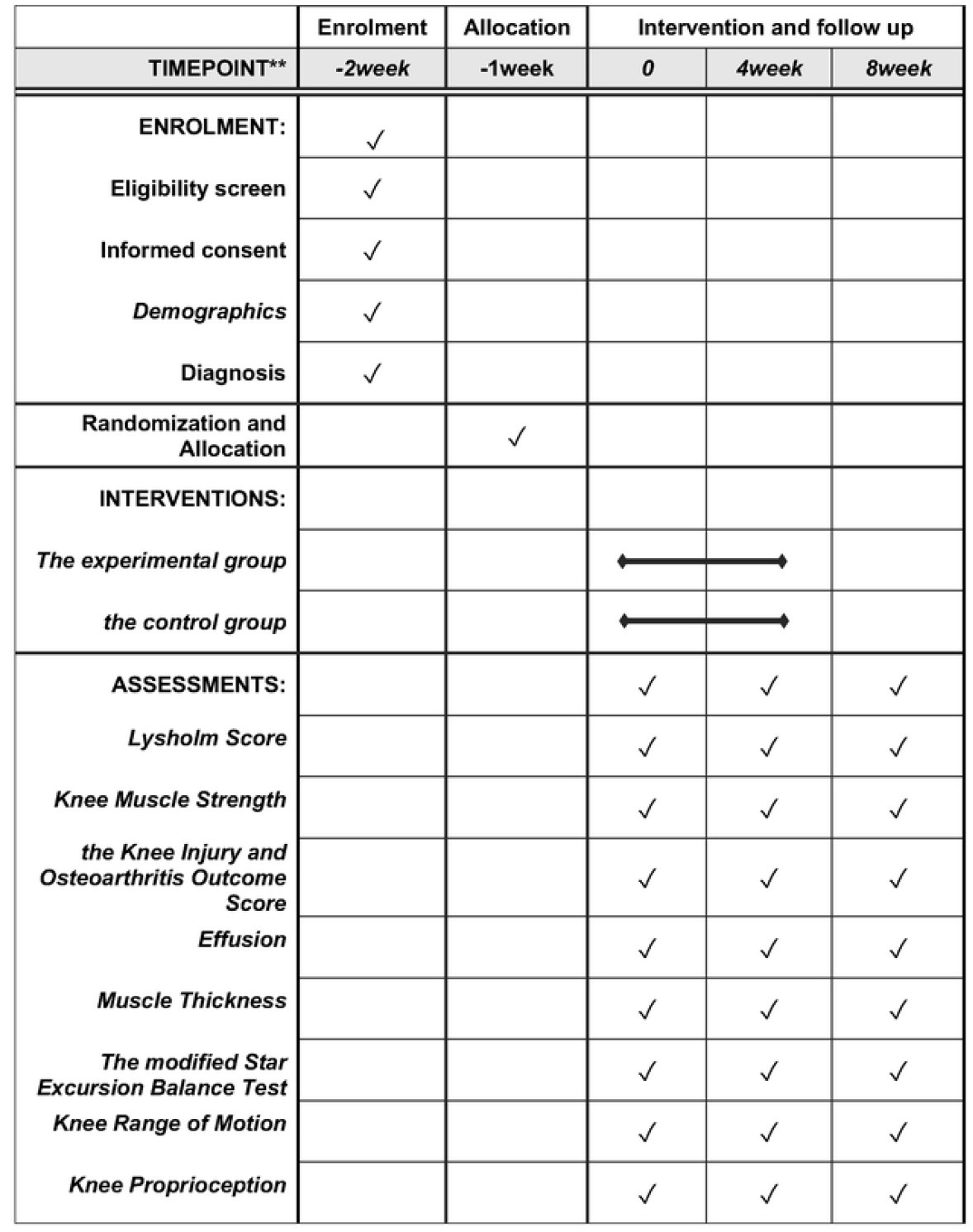
Intervention schedules.

